# ESSENCE, the Electronic Surveillance System for the Early Notification of Community-Based Epidemics

**DOI:** 10.1101/2020.08.14.20175398

**Authors:** Howard S. Burkom, Wayne A. Loschen, Richard A. Wojcik, Rekha S. Holtry, Monika A. Punjabi, Martina M. Siwek, Sheri H. Lewis

## Abstract

The Electronic Surveillance System for the Early Notification of Community-Based Epidemics (ESSENCE) is a secure web-based tool that enables health care practitioners to monitor health indicators of public health importance for detection and tracking of disease outbreaks, consequences of severe weather, and other events of concern. The ESSENCE concept began in an internally funded project at the Johns Hopkins University Applied Physics Laboratory (JHU/APL), advanced with funding from the State of Maryland, and broadened in 1999 as a collaboration with the Walter Reed Army Institute for Research. Versions of the system have been further developed by JHU/APL in multiple military and civilian programs for timely detection and tracking of health threats. Features of ESSENCE include spatial and temporal statistical alerting, custom querying, user-defined alert notifications, geographical mapping, remote data capture, and event communications. These features allow ESSENCE users to gather and organize the resulting wealth of information into a coherent view of population health status and communicate findings among users. The resulting broad utility, applicability and adaptability of this system led to adoption of ESSENCE by the Centers for Disease Control and Prevention (CDC), numerous state and local health departments, and the Department of Defense (DOD) both nationally and globally. With emerging high-consequence communicable diseases and other health conditions, the continued user-requirements-driven enhancements of ESSENCE demonstrate an adaptable disease surveillance capability focused on the everyday needs of public health. The challenge of a live system for widely distributed users with multiple different data sources and high throughput requirements has driven an novel, evolving architecture design.

## INTRODUCTION AND BACKGROUND

### Automated Routine Health Monitoring

Until recent decades, public health disease surveillance relied upon laboratory confirmation and passive participation. Often, the lack of automated detection and reporting resulted in time delays that impeded prompt mitigation activities. Public health institutions thus began using enhanced surveillance techniques with the potential for more timely epidemic detection and tracking. These techniques were incorporated in electronic and increasingly internet-based biosurveillance systems for everyday use by health monitors.

### Origin and Early Development

Intensive efforts to establish biosurveillance systems occurred at multiple institutions and government agencies in the late 1990s. The ESSENCE system originated as a collaboration of two such projects. A project of Dr. Michael Lewis, then a medical resident under Dr. Julie Pavlin at the Walter Reed Army Institute for Research, applied US military clinic visit data for outbreak detection in a project called “ESSENCE”, Electronic Surveillance System for the Early Notification of Community-based Epidemics (1). Concurrently, JHU/APL was combining civilian data from hospital emergency departments, physician office visits, over-the-counter (OTC) sales, and school absenteeism records, first with internal funding, then for the State of Marylance(2). The two groups joined forces in anticipation of possible bioterrorist activity at the turn of the century as January 1, 2000 approached, and this collaboration along with similar efforts at other institutions led to further funded development in the multi-center Bio-Event Advanced Leading Indicator Recognition Technology (BIOALIRT) program of 2001-3 for the Defense Advanced Research Project Agency (DARPA) (3). In leading one of four BIOALIRT research teams, the JHU/APL and WRAIR groups further matured the ESSENCE system concept.

> Following the terror attacks of September 11, 2001, ESSENCE was expanded in Maryland, increasingly operationalized in state and local civilian health-monitoring agencies, and implemented in all global US military treatment facilities. Since then, fueled by a succession of initiatives driven by both bioterrorism and natural public health concerns, versions of ESSENCE have been implemented in the Department of Defense (DoD), the Veterans Administration (VA), the Centers for Disease Control and Prevention (CDC), and state and regional public health agencies across the United States and, through collaborative DoD efforts, internationally. Following widespread use of ESSENCE and enhancements at JHU/APL to meet users’ evolving needs, the CDC National Syndromic Surveillance Program adopted ESSENCE in 2014 as the standard analytic surveillance and visualization engine on the BioSense platform for state and local public health monitors (4, 5). User benefits of ESSENCE have long surpassed the “Early Notification” part of its acronym to include aspects of situational awareness such as tracking and characterization of known health events, assessing the burden of all-hazards threats such as severe weather, environmental hazards, and substance abuse, and rumor control to enable improved public health response.

### ESSENCE History Highlights

1997: Early research in hospital information management led to a request by Maryland’s Secretary of Health to work with the Maryland Department of Health and Mental Hygiene to develop an automated system to detect bioterrorist attacks.
1999: The precursor to the current ESSENCE system, the Maryland Disease Surveillance and Reporting System (MDSRS), was implemented to monitor for bioterrorism. Subsequent collaboration with the Department of Defense Global Emerging Infection System led to DARPA’s multi-center BIOALIRT program enabling improvements in algorithmic detection and biosurveillance system architectures.
2001: ESSENCE moved from a dynamic web framework to the static pre-built visualizations to exploit commercial GIS technologies. Limitations on user capabilities forced a rapid return to the dynamic web framework with a database backend for ESSENCE II. Soon after the terrorist attacks of September 11, ESSENCE II was transformed from a limited-use research project to a live operational civilian surveillance system in Maryland and for all global US military treatment facilities.
2003: In response to the need to monitor the health of deployed US troops in the Iraq War, ESSENCE III was developed and deployed in March 2003. This version featured novel data sources, customizable syndrome groupings, and the first incorporation of external detection algorithms. The ability to implement external analytic methods prompted and validated the addition of an Application Program Interface (API) to the analytics architecture.
2003: Global emergence of the Severe Acute Respiratory Syndrome (SARS) spurred the need to sharpen and narrow case definitions in place of general syndromes. This threat solidified the decision to include free-text chief complaint queries along with diagnosis codes in the aggregations of monitored counts and rates. Tracking of SARS highlighted the power of automated systems for situational awareness, a broader and far more frequently useful objective than novel event detection. Subsequent experience began the expansion of syndromic surveillance from monitoring broad infectious syndrome groups to ad hoc categories of greater clinical specificity and a broader variety of population health threats.
2004: Expansion: the DoD Joint Services Installation Pilot Project (JSIPP) and the Department of Homeland Security (DHS) BioWatch program proliferated ESSENCE instances across the US. The influx of new users and widespread user sentiment that shareable and explainable results are no less challenging and important than event detection drove enhancements in visualization flexibility and exportability.
2004: The states of Maryland and Virginia and the District of Columbia agreed to share data across state lines, and the “Aggregated National Capital Region” (ANCR) version of ESSENCE was created. The Enhanced Surveillance Operating Group (ESOG) of ANCR users was formed to help guide ESSENCE development. The close tie between users and developers became a foundation for future ESSENCE enhancements.
2006: Expanded usage of ESSENCE including patient data from hundreds of hospitals at the Florida Department of Health led to additions to analytics including “time of arrival” monitoring in which anomalous groups of emergency department (ED) arrivals at hourly intervals were detected and reported with adjustments for patient volume, time of day, and subsyndrome type.
2007: The high-profile mass gathering events of the Super Bowl and the 2009 Presidential Inauguration impelled testing of information-sharing, distinct from data-sharing, strategies including an InfoShare tool to enable timely sharing by ANCR users with national level authorities. This effort foreshadowed multiple events or threats in which local restrictions prevented ESSENCE user sites from sharing explicit data, but derived reports, aggregates, data-free query language, or just descriptions could be legally shared. Implementation of ESSENCE features to facilitate such sharing has continued since then.
2009: Collaboration of the ESSENCE team with DoD-GEIS clarified the need to transition the technology and lessons learned from ESSENCE to an open source version that would be easy to operate, cost-free, and deployable around the world. The Suite for Automated Global Electronic bioSurveillance (SAGES) toolkit was developed, including OpenESSENCE and ESSENCE Desktop editions. While based on the features and functionality of ESSENCE, tools within SAGES were developed specifically for use in low- and middle-income countries. Working with the Veterans Affairs and DoD, the ESSENCE team investigated the benefits and obstacles of including elements of electronic medical records beyond the demographics and chief complaints. Working with multi-terabyte database sites prepared ESSENCE developers to deal with large datasets in every jurisdiction.
2013: The Boston Marathon bombings led to ESSENCE application to monitor non-infectious health categories including Anxiety, Depression, Suicidal Tendencies and Hearing Loss. These additions helped track the health of the Boston communities in the aftermath of the attacks.
2014: CDC announced the transition of the National Syndromic Surveillance Program (NSSP) to ESSENCE for syndromic systems analytics, to be hosted on the cloud-based BioSense Platform.
2016: Non-syndromic alerting algorithms were added to ESSENCE for rare, nonmedical, or uncommonly frequent terms in hospital free-text chief complaints.
2017: The opioid overdose crisis led to new, sustained partnerships between ESSENCE users and other public health divisions, such as injury prevention, behavioral health, and drug abuse. Analytic methods were explored to combine multiple data types for improved situational awareness. The MyESSENCE feature was added to allow users to customize dashboards for concise daily monitoring.
2018: The NSSP Syndrome Definition Committee users began sharing combined free-text and diagnosis code-based definitions for inclusion in local ESSENCE sites. Advanced natural language processing methods such as word embeddings were developed and shared among CDC and local users
2019: Nationwide monitoring of outbreaks of lung injuries associated with the use of vaping products increased demand for and development of system features to assess current data quality, including availability and informative content of the records underlying ESSENCE data displays.
2020: Collaboration among NSSP ESSENCE user groups intensified with evolution and sharing of queries related to COVID-19 pandemic surveillance.

## SYSTEM DESCRIPTION AND PRINCIPAL FEATURES

### System Overview

ESSENCE is an enhanced health surveillance system using advanced mathematical/visual analytics which can detect anomalies in both traditional and non-traditional public health data, with the goal of enabling public health to find and monitor outbreaks of health events and make decisions. Users of ESSENCE access a secure web-based tool to conduct disease surveillance for the purpose of timely detection, situational awareness, and descriptive epidemiologic analysis of baseline disease patterns and outbreaks. For effective public health response, public health authorities must have the ability to identify the infected population so further spread can be contained. Leveraging the near real-time availability of an increasing number of data sources, ESSENCE analytical and alerting capabilities provide an opportunity for public health users to capture the early stages of an outbreak and track its progress. ESSENCE enables integration of electronic data from both clinical and nonclinical sources to enhance situational awareness. For most users, the primary clinical data source is hospital emergency department chief complaint records. Additionally, based on availability, public health agencies incorporate other data types such as over-the-counter medication sales, poison control call center data, prescription drug data, reportable disease data, vital statistics mortality data, and school absentee data. Once raw data reach ESSENCE, the analysis, visualization, and communication features of ESSENCE allow the end user to gather and organize the resulting wealth of information into a coherent view of population health status and to communicate findings among other users and stakeholders.

As a result of ongoing user feedback daily and event-based disease surveillance needs, ESSENCE features include spatial and temporal statistical alerting, custom querying, user-defined alert notifications, geographical mapping, remote data capture, and event communications.

### Main Functions

ESSENCE provides three main functions:

1. *Data ingestion/preprocessing*. Traditional and non-traditional data sources are electronically received by ESSENCE and many are mapped to syndrome groupings. During the ingestion process, data are cleansed (e.g. duplicate records removed, invalid characters removed, etc.), updated (e.g. existing records information is updated with new information) and categorized (e.g. syndrome and subsyndrome categorization assigned to records).
2. *Alerting*. Multiple temporal and spatial alerting algorithms are applied to each data set to develop a list of alerts or flags for further investigation by public health officials. In addition to algorithms developed by JHU/APL and Walter Reed Army Institute of Research (WRAIR), ESSENCE can incorporate algorithms required by the jurisdiction where the system is deployed.
3. *Analysis and Visualization*, ESSENCE data and alerts can be analyzed and visualized in multiple ways in the system, both spatially and temporally.

Section 4 describes these functions in three subsections: Data Management, Alerting Algorithms, and Visualization.

## TECHNICAL DETAILS

### Data Management

#### Architecture

The software architecture employed for ESSENCE is a three-tier web application with a presentation layer as a user frontend, a business layer for application of algorithms, and a backend for databases. This architecture runs on modular server configurations, with the number of servers contingent upon the data volume, number of active users, and frequency of required analysis operations. The most common configurations comprise three servers for smaller instances and five servers for larger ones. For systems with larger numbers of data sources and/or data volumes reaching billions of records, the architecture can support additional servers to spread the functional load of the processing. The backend databases are Microsoft SQL Server relational database management systems (RDMS). Database functions include an ingestion database layer that facilitates extract-transform-load (ETL) operations and performs deduplication and other data cleaning operations. Depending on the user site, the ETL operations are performed by Rhapsody,(6) Mirth,(7) or locally developed scripts to populate the database, and then ESSENCE’s Groovy-based data flow management system (8) controls data flow and business logic to transfer data from ingestion to detection to web databases.

A detection database layer holds data and manages cube tables for fast algorithm access and execution. Java-coded algorithms access the data and cubes for efficient signal detection on the detection database to separate algorithm processing from user query management. The web database layer expedites rapid formation and display of interactive screens for visualization and communication. Web applications encoded in Java and JavaScript utilize this database via a Tomcat web application server.(9) For display purposes, mapping and other geographic information system (GIS) operations employ the open source tool GeoServer.(10) Users can access the web application through standard web application displays or via a web service API layer for direct access to ESSENCE data and functionality.

#### Data Types

The types of data analyzed in ESSENCE are the prerogative and responsibility of the jurisdiction, though JHU/APL provides capability for basic types. The ESSENCE system is data agnostic—the only requirement for a monitored data type is the inclusion of a data field. Data time resolution is also unrestricted. Data frequencies in ESSENCE have ranged from seconds to years, though daily data have been most common. All but a few users monitor hospital emergency department (ED) data. Most ESSENCE user jurisdictions face the burden of acquiring their data sources, gaining approval for their routine intended use, and extracting features to monitor. That level of effort varies greatly by data source. Various users also monitor or have monitored records of the data sources listed below.

- over-the-counter remedy sales
- physician office visits
- laboratory test orders and results
- school absenteeism and health office/nurse records
- reportable disease cases
- poison center call records
- ASPR DMAT data for disaster response (11)
- cardiovascular and other chronic diseases from inpatient encounters
- livestock and companion animal health encounters
- human vitals measurements
- death records
- emergency medical services (EMS) and 911 calls, and
- climate and air quality data

Users have also incorporated or adapted data from other surveillance systems such as the National Poison Data System (NPDS).(12) While not all of these sources have proven useful for routine surveillance, users have employed the system to investigate their utility and to seek best data usage practices.

In separate studies or projects, individual user jurisdictions or their research partners have also used ESSENCE to analyze records of radiology impressions, genomic sequencing data, zoo animal health, environmental sensor outputs, sales of specific products such as thermometers, orange juice, and tissues, social media posts and searches, and even fantasy sports data. Multiple content formats used in ESSENCE have included Health Level-7 (HL7) formats for hospital data, National Emergency Medical Services Information System (NEMSIS) formats for EMS data, and others. File formats have included delimited/tabbed/fixed-width American Standard Code for Information Interchange (ASCII) text, extensible markup language (XML) and JavaScript Object Notation (JSON).

#### Data Security

All automated data transfers occur over a secure virtual private networks (VPNs), e.g. via secure file transport protocol (SFTP); or over VPN tunnels, e.g. HL7; or by web transfer from a secure website, e.g. NPDS and weather. The use of web application access control allows limited, hierarchical access rights for every user by data source, by fields within a data source (e.g. geography or jurisdiction, syndrome), and by website function (e.g. data details, time series, alerts). These access privileges are customizable in the sense that one user site may control access by health district, others by county or by neighborhood. Some sites build aggregated datasets that meet the monitoring needs of users with limited access rights. Weather and other data sources with no privacy concerns may be made available to all users.

#### Data Preprocessing and Quality Management

Multiple, data-dependent preprocessing steps include deduplication procedures, formation of syndrome fields, calculation of distances, and deriving additional fields and flags based on jurisdictional business rules and logic.

Procedures for managing data quality issues such as deduplication, temporary dropouts of data feeds, data field value validation, and management of free-text or pick-list entries are incorporated in ESSENCE standard business rules. By these rules, ESSENCE does not update records in place but employs a delete-and-replace approach that has proven faster. For data with the functionality enabled, a history system integrates all prior instances of each record to produce a single master record with the relevant fields for each encounter. An extensive set of reference tables and business logic allows conversion of field entries such as patient age, race, ethnicity, and vitals measurement such as temperature to categorical values from standard sources such as the Public Health Information Network Vocabulary Access and Distribution System (PHIN VADS). A “region” data field is used for general spatial aggregation of patient records and is most often employed to combine count data from collections of zip codes to approximate county-level counts when the county field is unavailable. Beyond these features and conventions, ESSENCE includes a substantial website section with guidance and analysis tools dedicated to helping users manage the quality of their data.

### Alerting Methods using Statistics and Artificial Intelligence

#### Machine Learning and Natural Language Processing for Trackable Data Features

Data sources used or considered for health surveillance include medical encounter billing records, emergency service calls, nurse hotline calls, prescription and over-the-counter remedy sales, absenteeism records, and more recently, social media data such as tweets and web searches. Each data source has its own challenges for user jurisdictions to obtain sustained electronic access from data providers and any requisite government approval. When a data stream of any of these sources is acquired for routine monitoring, an immediate question is how to use the streaming data to track health outcomes of concern. An often applied procedure is to track counts of subcategories of the data expected to correspond to these outcomes. These subcategories are commonly called *syndromes*, generalizing the medical definition of this term denoting disease-related collections of signs and symptoms. Thus, in the surveillance context a *syndrome* may refer to grouped hospital visits associated with a fixed collection of symptoms, laboratory tests ordered for certain conditions, web searches containing sets of terms, billing records covering any of a class of remedies, or other subgroups depending on the data source. Syndrome formation is a critical step that may use only a fraction of all streaming data and may produce few or many groups to track. The number and composition of syndromes depends on the richness of the data, the number of outcomes of interest, and the resources of the monitoring institution for investigation and response.

Syndromes and subsyndromes used in ESSENCE vary depending on the clinical grouping systems available and the needs of the user site. Early versions of ESSENCE formed syndrome groups using diagnosis codes, which have disadvantages of late assignment and emphasis on billing practice in many medical systems. Examples were Respiratory and Gastrointestinal with subsyndromes such as Asthma. Categorization soon switched to the use of free-text chief complaint (CC) or reason-for-visit data fields. For this categorization, the JHU/APL ESSENCE team developed the Chief Complaint Processor (CCP), a versatile, stand-alone program for weighted keyword-based classification by free-text fields. The CCP is highly configurable, with tables including sets of syndromes and subsyndromes with classification rules allowing complex logic, positive and negative weighting of component terms, abbreviation and spelling rules, and a list of unmodifiable terms. For example, CCP puts a record with a CC of “NAUSEA” or “VOMITING” in the Gastrointestinal (GI) category. The CCP creates a ChiefComplaintsParsed field for use of classification rules after treatment of abbreviations, some misspellings, and other cleanup.(13) These classifications have enabled additional natural language processing and machine learning initiatives by both ESSENCE developers and users, and findings from these initiatives are shared among users with each emerging health threat.(14–16)

As done with diagnosis code-based processing, syndrome groups are tabulated, plotted and monitored each day with statistical alerting algorithms for early potential outbreak indications.

#### Application Principles for Alerting Algorithms

Individual alerting algorithms implemented in ESSENCE are listed and described in the Supportint Materials file “S1 Technical_Details_of_ESSENCE_Alerting_Algorithms.docx”.

The following principles were derived with users to guide method selection and to clarify interpretation of results:

General considerations:

- These methods are not intended to positively identify outbreaks without supporting evidence. Their purpose is to direct the attention of a limited monitoring staff with increasingly complex data streams to data features that merit further investigation. They have also been useful for corroboration of clinical suspicions, rumor control, tracking of known or suspected outbreaks, monitoring of special events and health effects of severe weather, and other locally important aspects of situational awareness. Successful users value these methods more for the latter purposes and do not base public health responses solely on algorithm alerts.
- These algorithms are one-sided tests that monitor only for unusually high counts, not low ones. Low counts could result from a critical outbreak situation that prevents data reporting, but there are many more common reasons for low counts (such as unscheduled closings or system problems), so the algorithms do not test for abnormally low counts.
- In addition to data- and disease-specific considerations below, algorithm selection was also driven by system considerations. Users need to monitor many types of data rapidly. External covariates such as climate data or clinic schedules may not be available for prompt analysis. Many methods in the literature, armed with substantial retrospective data of a certain type, depend on analysis of substantial history. Day-to-day users, often with only a small fraction of time available for monitoring, will not wait several minutes for each query. In the absence of data history and data-specific analysis time for each stream, ESSENCE methods have been adapted from the literature and engineered to system requirements.
- If the time series monitored by algorithms represent many combinations of clinical groupings, age groups, and geographic regions, excessive alerting may occur simply because of the number of tests applied. The Summary Alert method was implemented to limit such excessive alerting. This method is based on control of the false discovery rate, i.e. the expected ratio of false alerts to the total alert count, and its statistical implementation in ESSENCE is detailed in the Summary Alert section below. Aside from analytic methods to control alerting, default alert lists should be limited to results from those time series of concern to the user, either by system design or by active specification by the user. For example, one method of reducing the default alert list is to restrict algorithms to all-age time series groupings. Depending on the scope of the user’s responsibility, the alert list may also be restricted according to both epidemiological interest and the resources available for investigation. For example, a monitor of a national-level system with algorithms applied to many facilities may be interested only in alerts with at least 5-10 cases. In circumstances of heightened concern, these restrictions can be relaxed, or the user can use ESSENCE advanced querying methods to apply algorithms to age groups and/or subsyndromes.

#### Visualization

Both standard and user-customizable visualizations are available in ESSENCE. Highly configurable and interactive modes of data stratification and filtering, graphical and tabular customization, user preference management, and sharing features allow users to query data and view geographic representations, time series and data details pages, and reports. The following sections summarize key features. Screenshots of selected novel visualizations are shown below.

#### Standard Visualizations

Following are the most commonly used ESSENCE visualizations:

- The Time Series View provides a graphical display of the temporal behavior of the data with the ability to stratify by specific parameters, view aggregated counts, and infuse data quality factors to improve understanding of data features.
- The Data Details Page provides line listings of individual records and pie/bar chart representations of query results.
- The Map View allows the user to view both data and alerts on a geographical display of the specified region.
- The Alert List provides a view of signals generated by the alerting algorithms. Each row of the list includes a link to the corresponding Time Series View or Data Details Page.

To facilitate selection of desired data by space, time, subpopulation, and clinical specificity, the Query Portal provides the user with tools for detailed design and management of simple or complex queries for routine or ad hoc monitoring. An online Query Wizard facilitates these processes as in Fig 1. For the desired analysis, the user chooses the data source, date range, time resolution (daily, weekly, monthly, quarterly, or yearly), temporal detection algorithm, and selections from a variety of component filters depending on the data source. For each data source, the user can define custom filters through configuration files that customize visualizations. Options include Free-Text, Reference List, Number Range, and Dates filters. Examples of query field options for filter creation are age group, geography system, syndrome or medical category of interest.

**Fig 1:**
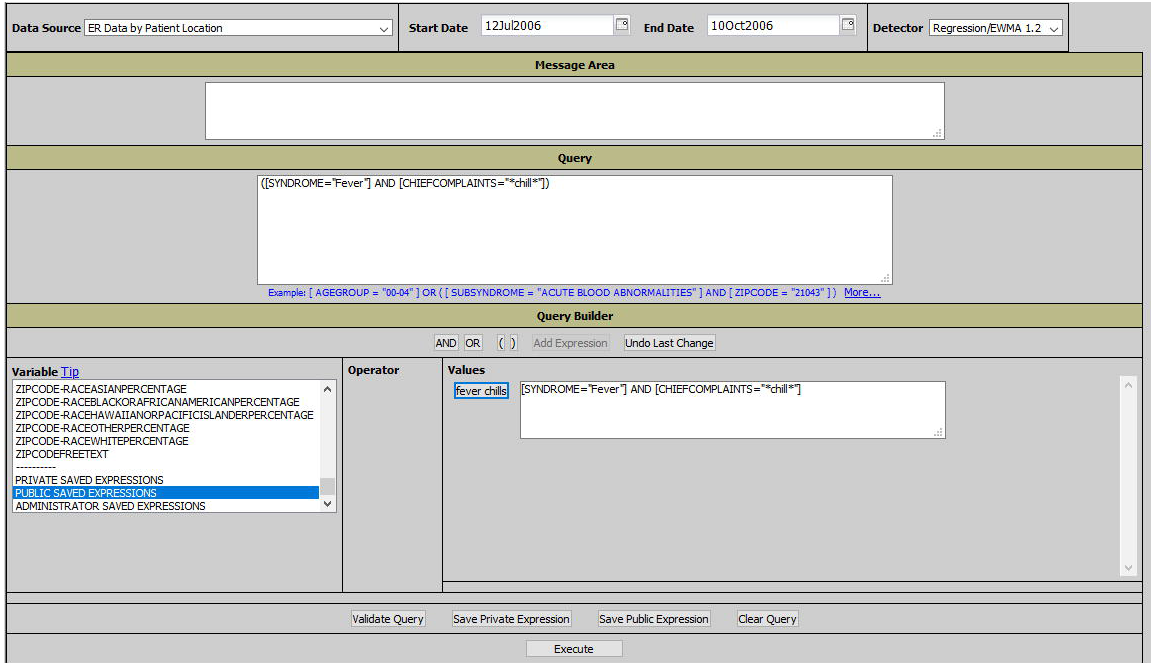
The ESSENCE Query Wizard.

Details of the attributes available for selection, stratification, and filtering are:

- **Geography System:** Data can be viewed in many geography systems such as Region, Zip Code, Hospital, Region of the Hospital, Military Treatment Facility (MTF), School, or Store. Each geography system defines a system for geographically filtering your data. Regions are a generic term that defines the default geographic way to view data. Regions normally map to a set of zip codes that closely resemble a county or health district.
- **Medical Grouping System:** When viewing the data via the Query Portal, the user has the ability to choose between how the various data are presented. Data may be viewed by many grouping systems, including ESSENCE Syndrome, International Classification of Diseases (ICD) Code, Chief Complaint SubSyndrome, Chief Complaint, OTC Category for records of sales of over-the-counter remedies, or Call Center Guideline, depending on which data source is queried.
- **Syndrome Grouping:** The syndrome groupings used in the ESSENCE system vary depending on the medical grouping systems used and the needs of the user site. These groupings are used to filter data into medically similar sets. Examples of a syndrome grouping are: Respiratory, Rash, Cough/Cold, Sinus, Asthma, Chest Pain, etc. Each medical grouping system will have a set of syndromes or the ability to perform free text queries. A Syndrome Definitions user interface provides a stepwise mechanism for viewing rules that define a syndrome or subsyndrome. In addition to syndrome categories the user may also query based on a Chief Complaint Discharge Diagnosis (CCDD) field, a concatenation parsed chief complaint and discharge diagnosis. Filtering by CCDD category uses Structured Query Language (SQL) “Where” clauses to select records meeting user criteria. In general, this filtering takes the form of simple keyword matching with inclusion of wildcard matching and negation terms.
- **Detector:** The ESSENCE tools refer to alerting algorithms as *detector*s. For temporal alerting methods, users may choose any of the methods described in Section 4.2.1.

For queries involving more complex data selection and free-text logic than the query wizard provides, ESSENCE provides an Advanced Query Tool, shown in Fig 2.

**Fig 2:**
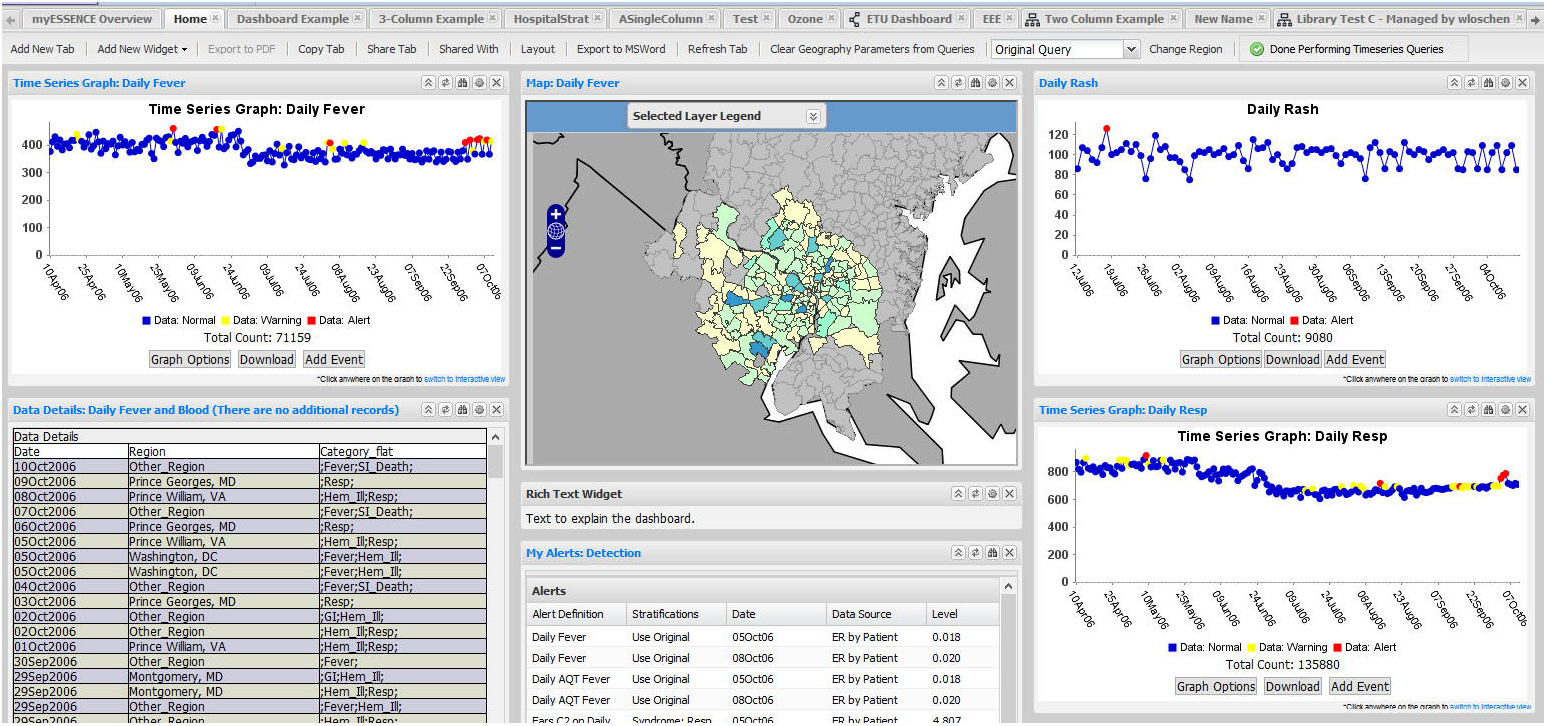
The Advanced Query Tool in ESSENCE.

Once a user has created a query, an action button is available to indicate the use and disposition of the query. Options include formation of time series and tables. Complex queries can be saved for reuse and for application to other appropriate data sources.

#### User-Customizable Visualizations

In addition to the standard visualizations, ESSENCE offers additional views and analysis modes. Of these, the most commonly used are myESSENCE and myAlerts.

##### myESSENCE

The myESSENCE feature on the main ESSENCE menu facilitates creation of multiple customized dashboards of graphs, charts, tables, maps, and alerts. Users may create separate dashboards by using a widget interface to select, drag, and drop widgets. They may share each dashboard with other users by providing a copy to the dashboard or by providing a read-only copy over which they maintain control. Possible choices for the widgets are time series graphs, maps, and listings of data details. Parameters for each view, such as start and stop dates, are modifiable. These views may be arranged in 1-, 2-, or 3-column format. Fig 3 exemplifies the dashboard creation process.

**Fig 3:**
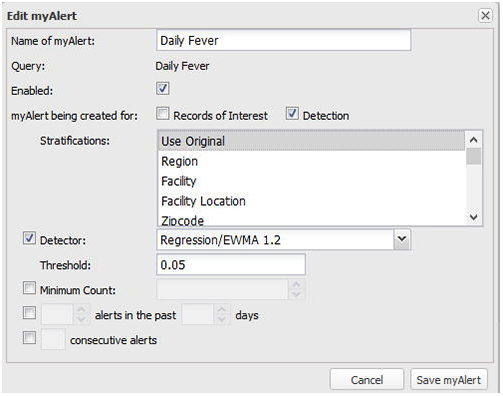
Formation of Customized Dashboard Using MyESSENCE Feature.

The creating user gives each dashboard a text description and a note that is modifiable by creator or shared user. Once created, each dashboard appears as a separate tab on the myESSENCE webpage.

By default, each dashboard applies to data from the geographic regions selected for each graph when added to the dashboard. The creator or sharing users may change the region, and ESSENCE will change all views on the dashboard to reflect data from the new region. Users may revert to the default view by choosing “Original Query”.

##### myAlerts

The myAlerts feature enables users to set up automatic notifications of query results reflecting specific subpopulations, algorithms, and conditions. Two types of these notifications are available. Notifications triggered by receipt in ESSENCE of data records that satisfy a query definition are *Records of Interest* alerts. Notifications triggered by minimum count requirements or by algorithms detecting sets of records returned by a query are *Detector* alerts. Algorithms specified may be any of the methods described in Section 4. The user may also specify a minimum number of records requirement in addition to an alerting algorithm. Fig 4 shows the web page used to create this customized alerting feature. Alerts that require algorithm results that cross a designated statistical threshold may be restricted to require threshold crossings on m consecutive days, or on m of the last n consecutive days. The user may set up automatic email prompts for each customized alert and may choose automated sharing of selected alerts with other designated ESSENCE users.

**Fig 4:**
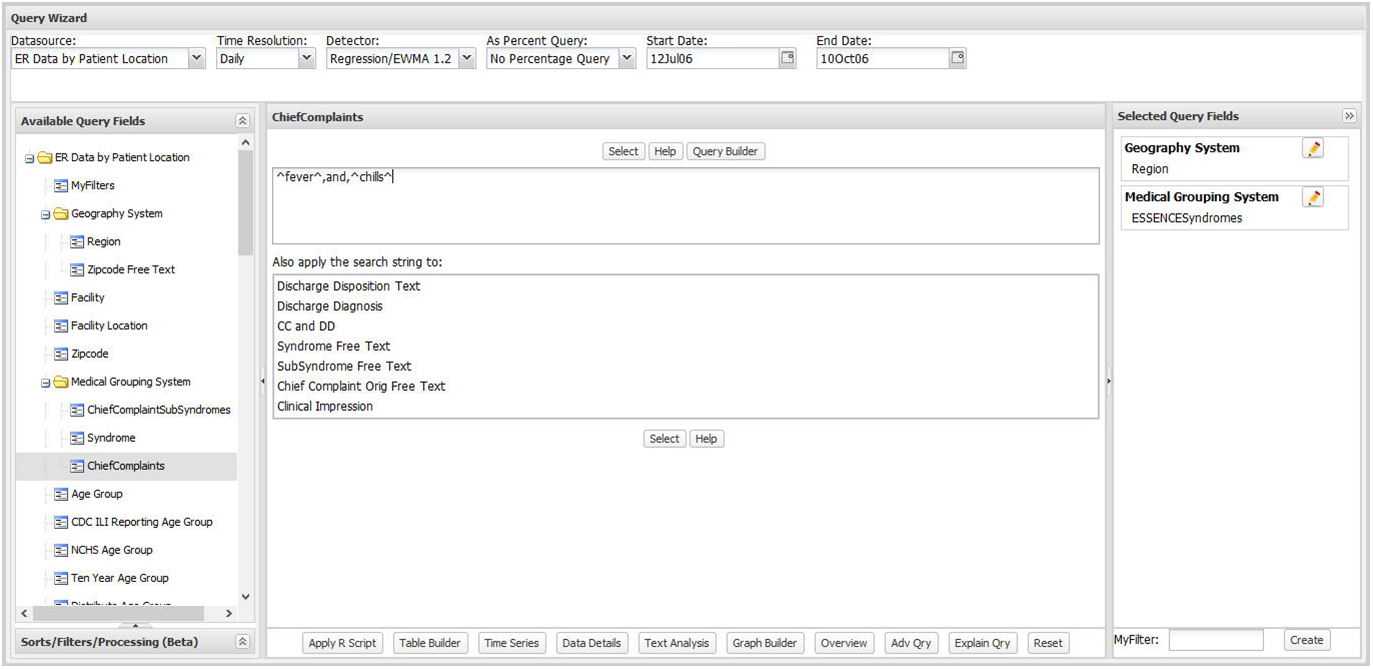
Creation of MyAlert for Customized Anomaly Detection.

Developers of ESSENCE have also created single-purpose views and tools for specific jurisdiction needs, including statistics tables and side-by-side graphs, as broadly applicable as possible to all data sources.

## USER APPLICATIONS AND SUCCESSES

This section describes ESSENCE applications in health departments for analysis of a wide range of public health threats in monitored populations.

### Infectious disease applications

Early applications of ESSENCE focused on detection of infectious disease outbreaks, with much attention on influenza-like illness (ILI) because ILI is a prodrome for multiple naturally occurring diseases and for many potentially weaponized for bioterrorism (35, 36). Civilian (37), military (36), and VA (38) ESSENCE users monitored for outbreaks of seasonal and nonseasonal outbreaks of febrile respiratory infections, GI infections caused by contaminated food or water, and rarer infections.

Surveillance systems were particularly helpful to track the pandemic of novel H1N1 influenza in 2009 (38). The pandemic was important as an interregional use of surveillance systems to track a common threat. For example, the National Capital Region (NCR) Disease Surveillance Network, comprising ESSENCE users at health departments in the District of Columbia and parts of the states of Maryland and Virginia, shares population-level disease incidence information to promote inter-jurisdictional surveillance. In 2009, this network allowed NCR public health practitioners to track the course of the pandemic from the spring through the fall, comparing the overall and age-specific burden of illness to national and neighboring state trends. A broader collaboration of both ESSENCE and non-ESSENCE system users collaborated on a standardized definition of ILI to enable uniform local tracking of the pandemic across the US.(39)

Users have applied ESSENCE to form and share queries for indications of other infectious diseases using text from chief complaints, discharge diagnosis, and triage notes when available. Infectious threats tracked in published examples include general waterborne diseases (40), tuberculosis (16), rabies (41), and Middle East Respiratory Syndrome (14). The recent years’ concerns over mosquito-borne diseases has also generated new queries by multiple users and occasionally uncovered important cases.(15)

The health department of Maricopa County, Arizona presented an example of the benefit of surveillance systems in June, 2018. The department had added an ESSENCE query for signs of Rocky Mountain spotted fever, which is not endemic to that county, because of concerns that cases transferred from endemic areas might be missed. A child’s patient record was signaled by the query, and the department contacted the hospital. This contact led to reversal of a medication decision that might have been fatal to the child.(42)

Such monitoring activity has repeatedly uncovered unreported cases of diseases for which reporting is mandatory. (43) These findings illustrate the importance of redundancy with systems such as ESSENCE to avoid missing important cases, even when traditional reporting mandates exist.

At submission of this manuscript, intense collaboration of ESSENCE users is focused on tracking of the COVID-19 pandemic. General queries on COVID-like illness and specific ones involving pneumonia and specific symptom sets are being refined and shared.(44)

### Applications for tracking burden of severe weather, natural disaster events

Health departments have used ESSENCE for preparedness, health burden assessment, and response to severe storms and other natural disasters. The state of Oregon conducted a successful program to mitigate the effects of wildfires.(45) This program featured customized ESSENCE queries with other coordinated efforts among state and local health departments and preparedness teams. Among several states that using ESSENCE to monitor the effects of hurricanes, the Tennessee health department devised queries to determine the volume and clustering of patients in local hospitals because of storms in other states.(46) Effective monitoring of some events requires combination of multiple data sources. Following a prolonged storm-related power outage, the health department of Seattle-King County, Washington combined data from ESSENCE emergency department data with ambulance call and public utility data to monitor for cases and clusters of carbon monoxide (CO) poisoning and food poisoning.(47) More recently, the Florida health department monitored for CO poisoning after Hurricane Irma in 2017.(48) Institutions using ESSENCE are increasingly incorporating environmental and other data sources in their systems for richer situational awareness of disaster-related health threats. (49, 50)

### Applications for mass gathering surveillance

Scheduled mass gathering events such as political conventions and major athletic competitions concern population health monitors because a) such events are bioterrorism opportunities to affect many victims and gain media attention, b) infections through contaminated food or water could spread rapidly through the expanded population, c) those visiting for several days could import infections or take them back to their own cities, and d) a surge of patients could overwhelm local care provider resources. Adequate preparedness and response require coordination across jurisdiction boundaries, but privacy laws often restrict patient-level data-sharing. ESSENCE syndrome definitions and queries have been customized for many such events. In 2007, Marion County, Indiana and Cook County, Illinois were home counties for the competing teams in Super Bowl XLI, and the game was hosted in Miami-Dade, Florida. Health departments of these geographically distant counties were ESSENCE users, and customization of their systems for the days surrounding the event helped coordinate surveillance despite only two weeks’ notice after teams were determined.(51)

A partnership of the Florida Department of Health and the U.S. HHS Office of the Assistant Secretary for Preparedness and Response (ASPR) to improve response by Disaster Medical Assistance Teams (DMAT) produced a new ESSENCE module that was deployed for health monitoring of the 2012 Republican National Convention in Tampa.(50) State and county health departments used ESSENCE for coordinated monitoring of crowds at the US Olympic Trials in July 2016.(52) In January 2017, the Washington DC Department of Health used ESSENCE queries along with other data sources for health surveillance at the 58th US presidential inauguration.(53)

For monitoring events outside the US, JHU/APL used the SAGES system in 2014 to monitor the 8th Micronesian Games held in Pohnpei Federated States of Micronesia and the 3rd International Conference on Small Island Developing States in Apia, Samoa.(54)

### Applications for chronic disease and mental health surveillance

Usage of ESSENCE to monitor risk factors and incidence of chronic disease and mental health disorders has proliferated since a DoD ESSENCE study utilizing clinic and prescription data to monitor behavioral health in 2004. (55) The Boulder County, Colorado Health Department recently implemented and tested multiple queries to monitor mental health.(56)

Addressing ESSENCE utility for chronic diseases in general, the Cook County Illinois health department applied machine learning methods to assess the utility of ESSENCE emergency department (ED) data for monitoring cardiovascular disease (CVD), acute myocardial infarction (AMI), acute coronary syndrome, stable angina, stroke, diabetes, hypertension, asthma, and chronic obstructive pulmonary disease. From correlational validation testing based on eight full years of chief complaint text and electronic medical record data, they concluded that ESSENCE data are suitable for monitoring all of these conditions except stable angina and hypertension “at local, state, or national levels”.(57) The Nebraska state health department has used ESSENCE to monitor for CVD for several years, and the Florida state department similarly monitors AMI incidence.(58)

### Applications for injury and substance abuse surveillance

An unexpected but arguably the most helpful benefit of ESSENCE to health department users has been to facilitate communication and collaboration among agency divisions. An important example in the context of the ongoing opioid overdose crisis has been the strengthening of connections between syndromic surveillance specialists and groups specializing in injury prevention, behavioral health, and drug abuse.

Multiple health departments have applied ESSENCE to gain awareness of locations and subpopulations at risk for injuries from falls.(59) The Boston Public Health Commission in Massachusetts used it to monitor for hearing loss, acute depression, and explosion-related injuries in the aftermath of the 2013 Boston Marathon bombing and subsequent manhunt. The St. Louis, Missouri health department established ESSENCE queries for injuries indicative of bomb-making activities.(60)

Monitoring for substance abuse has been common among ESSENCE users. The Tri-County Health Department in Colorado uses its system to seek adverse effects of marijuana use.(61) The Florida state department queries for ED visits resulting from synthetic marijuana(62) and for novel street drugs such as Flakka as they become known public health problems.(63) Most recently, the opioid crisis has stimulated intense collaboration including shared syndrome definitions and analytic case-finding tools among geographically scattered institutions using ESSENCE.(64–66)

## CONCLUSIONS

### Lessons Learned

Several principles have driven the success of ESSENCE since its origins in the late 1990’s.

#### Versatility

Users have valued the configurability and adaptability of ESSENCE. Default categorization of complex data into syndromic groupings has always been valuable to users who are inexperienced or who do not have the time to formulate or validate their own categories for monitoring. Conversely, health departments with more analysis capacity have long demanded surveillance systems that let them create their own categories to track, and ESSENCE customization with query-building features using both diagnosis codes and free-text has grown along with the sophistication and broadening needs of health department users. Pre-computed, canned analysis products are not found in ESSENCE. However, versatility presents challenges to database design and to the selection and adaptation of statistical analysis tools. Surveillance data evolve with institutional information systems and formats, coding practices, and epidemiological concerns. Users typically cannot wait several minutes for data retrieval and time-consuming model runs. Alerting algorithms applied prospectively to detect disparate events in a wide variety of data types cannot match the detection performance of models developed retrospectively using historical datasets labelled with target events for a particular syndrome. Algorithm baselines in ESSENCE do not reach back for years, not only for storage and computational reasons, but also because for many users’ desired data types, stable data or any data are available only within the past year. Hence, ESSENCE alerting algorithms, adapted from published applications of models and control charts in healthcare settings, (23, 67, 68) use rolling baselines of weeks rather than years.

#### Facilitating communication

Multiple ESSENCE users have remarked that one of the system’s main benefits has been to facilitate communication with other divisions within a health department, with external local and federal agencies, and with care facilities that provide data and can benefit from the broader geographic perspective that a surveillance system enables. Hence, substantial ESSENCE development has occurred in response to user requests for custom analysis comparisons, visualizations, and report formats, allowing overburdened users to concentrate on the task of routine health monitoring. In situations where data-sharing is precluded by county or state regulations, ESSENCE communication tools have enabled information-sharing.

#### Multiple analysis modes

The applicability of individual analysis modes such as univariate and multivariate time series monitoring, spatiotemporal cluster detection, and single case identification all depend on the nature and quality of available data. For example, the spatial scan statistic implemented in ESSENCE can avoid issues of jurisdictional boundaries, but only if data location fields are present and reliably used in the data. In many data sources, the limitation of location fields to zip codes or postal codes restricts the geographic precision of clusters of interest. Health monitors generally need multiple ways to analyze population health data. The clearest example is that ESSENCE users in multiple health departments have discovered unreported cases of reportable disease that traditional sentinel surveillance is expected to communicate to public health. The various analysis modes of ESSENCE provide affordable and sometimes beneficial analytic redundancy.

### Innovation

The development of ESSENCE has produced novel features in areas of complex, disparate data management, analytical methods, and the enhancement of user reporting and collaboration, interrelated efforts to empower health monitors. The data management advances include architecture and data transfer capabilities to meet needs of institutions with varying resources. Analytics advances have required data quality examination methods and alerting algorithms appropriate for diverse data time series that meet rapid response needs and do not require more than a few months of data history. User experience enhancements have included customizable visualization and reporting features that provide unique time- and resource-saving capabilities.

### Impact

Recent ESSENCE projects have produced a variety of user capability enhancements. For sharing of information within and across jurisdictions, online features allow users to share what they are doing within ESSENCE with peers and to see what others are querying and find interesting. Text analysis and visualizations facilitate creation of ad hoc local free-text queries. These features provide correlation, trend, and association analytics to help the user determine what terms/phrases queries should or should not include.

Back-end tools and checks with visualizations allow the user to monitor closely the local ESSENCE system for data issues and irregularities. These administrative capabilities help managers and users maintain day-to-day system availability and improve visibility of issues that may develop over time.

Recently added visualizations and cohort clustering analytic tools for longitudinal assessment allow users to determine categories of patients who use healthcare systems that provide data to ESSENCE. These tools can show patient-level usage trends to inform allocation of healthcare resources in a community.

The ongoing adaptation of ESSENCE to meet the needs of the understaffed public health practice community has provided a means to share methods and information, though data are often not shareable. The common analytic platform has enabled an evolving user ecosystem of multiple working groups, and the US CDC currently hosts the NSSP Community of Practice, with subgroups including Syndrome Definition, Data Quality, and Technical committees.(69) The Syndrome Definitions committee promotes the analysis of common queries among geographically scattered user sites and US CDC,(70) thus improving communication between local and national health monitors. A primary example is the opioid overdose crisis, a noninfectious threat. In addition to analytics and visualizations to support the activities related to this crisis, developers and users have worked together to acquire additional data sources such as emergency medical incidents, poison center calls, and death records to determine the benefit of fusing information varying in specificity and timeliness into a common surveillance picture to better inform awareness and interventions.

### Future Directions: Challenges to ESSENCE and the Surveillance Community

Challenges to public health surveillance span multiple disciplines ranging from epidemiology and applied microbiology to statistics and machine learning and to database and network technology.(71) In ESSENCE, data processing/storage and analytics challenges go hand-in-hand. A major data challenge is to integrate increasingly diverse and granular data sources while preserving the essential features of ad hoc user syndrome/case definition and custom real-time analysis and visualization. Public health staff need to extend the power of their systems to all-hazards public health threats and to One Health issues including zoonotic diseases across species, antimicrobial stewardship, and food safety. Users of ESSENCE have long imported non-clinical and non-syndromic data sources such as pharmacy sales and school absentee rates along with traditional medical encounter records.

In expansion to address all-hazards threats, data complexity, categorization, and linkage challenges will multiply as genomic, environmental (including remote sensing) and social media data sources are added.

Significant advances in disease surveillance will also require meeting key analytics challenges. Beyond the monitoring of individual sources and syndromes with data dashboards, combining disparate data types requires statistical and machine learning tools for appropriate weighting and corroboration of evidence from disparate data types. Analytic fusion efforts employing Bayesian networks and other machine learning tools have been applied in both military and civilian ESSENCE systems (72–75). More efficient and explainable fusion methods will be needed to enable operationalizable forecasting and prediction for greater decision-making power and effective planning and response. Efficient analytic methods will also be needed to determine optimal feature extraction and resultant surveillance value of social media and other nontraditional sources.

Lastly, best practices for biosurveillance systems face both human and electronic communication challenges, including interoperability with other electronic systems such as the above-mentioned ASPR DMAT, NPDS, and NEMSIS systems. Despite the preeminence of ESSENCE, as indicated by its adoption as the analytic engine of the CDC Biosense platform and the widespread application discussed in this paper, the limitations described above and the data gaps exposed by the COVID-19 pandemic must be addressed as new threats emerge and data sources proliferate. These challenges must be addressed as ESSENCE matures to expand public health surveillance capabilities and utility beyond what is imaginable today and propel public health toward new frontiers of understanding and action.

## Data Availability

No data are required for this manuscript.

## ACKNOWLEDGEMENTS

We would like to acknowledge invaluable assistance from our colleagues and partners. The continued ESSENCE research and development would not have been possible without the support of the CDC, DoD, VA, numerous State and Local health departments,and JHU/APL over the last 20 years. We especially acknowledge numerous champions and advisors among ESSENCE users who have dedicated time and effort to the betterment of public health surveillance.

## Technical Details of ESSENCE Alerting Algorithms

### Univariate temporal alerting algorithms

The default temporal algorithm in ESSENCE is an automated selection between data modeling and control-chart-based algorithms, resorting to a simplistic Poisson distribution-based method if only a few days of recent data are available. The modeling method is adaptive multiple regression, while the control chart-like approach is an adaptive exponentially weighted moving average (EWMA) algorithm. The primary regression and EWMA methods are first discussed separately. Each description below gives a method category, purposes of the method, a brief technical description, key benefits, limitations, and literature sources.

### Algorithm: Linear Regression

#### Categorization

Adaptive Multiple Regression Model

#### Purposes

This model is an adaptive regression model applied to remove the systematic behavior often seen in time series of daily, syndromic, clinical visit counts and in other surveillance data. The reason for removing these common effects is to avoid bias in identifying unusual behavior. For example, there is a customary jump in visits on Mondays because many clinics resume normal hours, and this expected jump should not automatically increase the possibility of an alarm(17, 18). Similarly, alarms should be possible on weekends even though visit counts drop off from weekday levels.

#### Technical Details

This adaptive, multiple, least-squares regression algorithm contains terms to account for linear trends, day-of-week effects, and holidays. Multipliers for these terms are calculated using 4 weeks of recent counts as a training period. This training period is separated from the date of the test data by a 2-day buffer intended to keep early outbreak effects from contaminating the training. Extreme data values in the training period are reduced to reasonable values in order to avoid exaggerated predictions. This outlier correction for model inference avoids loss of sensitivity in the weeks after either data problems or true outbreaks. The regression multipliers are recomputed each day for calculation of a predicted count based on the expected data trends. The algorithm then subtracts this prediction from the observed visit count, scales the excess by the standard error of regression, and applies a statistical hypothesis test to determine whether to signal an alert. The test is a Student’s t distribution at significance levels of 1% for red alerts and 5% for yellow alerts, with the number of degrees of freedom determined by the number of regression covariates and the baseline length(19). Covariates and training intervals were chosen to obtain maximum sensivity for detection of injected signals at manageable background alert rates.

#### Benefits

The main benefit is avoiding alerting bias resulting from expected data trends. The length for the training baseline is critical. Based on performance comparisons among multiple baseline lengths, it was chosen to be short and recent enough to capture seasonal time series behavior but long enough to smooth out daily fluctuations. Separate multipliers are updated so that a data source with regular but unusual patterns such as high weekend counts will be modeled correctly. While a better fit may often be obtained with a more complex model for a given data stream with a certain syndromic filter for a certain subregion and analysis of sufficient data history, the current regression approach is relatively robust across time series employed in ESSENCE.

#### Limitations

If this algorithm is applied to a data series without the baseline weekly and seasonal behavior, the model will not explain the data well, and the detection sensitivity and specificity will be decreased. The automated switch in the default method is applied for this reason. There is no claim of optimal modeling for a given time series. This general-use implementation does not assume the availability of a denominator variable such as the total visit count that can be used to adjust the counts to emulate series of rates rather than counts. This adjustment has been implemented in particular versions of ESSENCE and in past versions of Biosense(20, 21) and could be added as an option.

### Algorithm:Adaptive Exponentially Weighted Moving Average (EWMA)

#### Categorization

Adaptive Control Chart

#### Purposes

This algorithm is appropriate for daily counts that do not have the characteristic features modeled in the regression algorithm. It is more applicable for Emergency Department data from certain hospital groups and for time series with small counts (daily average below 10) because of the limited case definition or chosen geographic region.(22)

#### Technical Details

This algorithm compares a weighted average of the most recent visit counts to a baseline expectation. For the weighted average to be tested, an exponential weighting gives the most influence to the most recent observations. Two weightings are applied: the first gives negligible weight to observations over 3 days old and is designed to detect sudden events where most outbreak cases affect data within a few days. The second weighting distributes influence further over the past week for sensitivity to more gradual outbreaks. These weightings emulate a dual strategy published for the hospital setting.(23) The monitored weighted averages are the Sk given by:

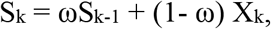

for a constant smoothing coefficient ω with 0 < ω < 1 and X_k_ as the successive data counts, with X_0_ = 0 and S_0_ = half the alerting threshold for prompt sensitivity. (Occasionally a useful starting value for X_0_ is known, but restarts may occur for many reasons, so the conservative initialization to 0 is used.) For separate monitoring of sudden and gradual events, smoothing coefficients ω = 0.9 and 0.4 are used. For both weighted averages, the 4-week baseline mean is subtracted, with a 2-day buffer period to separate the baseline from the counts being tested. The rationale for the baseline length was the same as described above for the regression method above. The test statistic is then (S_k_ – μ_k_) / σ_k_, where μ_k_ and σ_k_ are baseline mean and standard deviation. As in the regression method, the hypothesis applied to determine alerting is a Student’s t distribution at significance levels of 1% for red alerts and 5% for yellow alerts. The number of degrees of freedom assumed for this distribution is the baseline length + 1. This EWMA implementation is designed for any series that does not fit the characteristic trends, so a couple of safeguards are included. A “zero-filtration” algorithm is implemented for rapid adjustment to and recovery from data dropouts and catch-ups. When counts are sparse but not uniformly zero, a Poisson-based adjustment is added to the standard deviation scale factor to avoid excessive alerts.(19)

#### Benefits

This method gives sensitivity to both sudden and gradual outbreaks and has demonstrated prompt alerting capability. It is less susceptible than the Early Aberration Reporting System (EARS) methods C1, C2, and C3 to trends and to day-of-week effects. The added recovery features handle the most common problems in the data acquisition chain. Alerting is indirectly adjusted for the data distribution via the standardized residual test statistic, which provides a safeguard against excessive alerting when counts are small.

#### Limitations

This algorithm applied to pure daily counts does not control for expected trends or cyclic effects as in the regression method.

### Algorithm: Poisson/Regression/EWMA (default)

#### Categorization

Automated switch between data model and control chart

#### Purposes

Many researchers and developers have applied complex statistical models to surveillance data for prediction and detection. However, the predictive capability of a model varies according to the specific data stream and how it is filtered and aggregated. This capability may also be affected by data behavior changes that result from seasonal variations, population shifts, and changes in the informatics. To account for such day-to-day changes, ESSENCE automatically monitors its predictive capability of its regression model each day. When this test fails, indicating that the model is not helpful for explaining the data, the system switches to the EWMA adaptation described above. The result is that the regression model is usually applied for the common respiratory and gastrointestinal syndrome classifications applied to county-level data, but EWMA is more commonly applied to rare syndrome data. For situations where less than a week of recent baseline data exists, a simple Poisson detector is applied. Such situations include new start-ups and more common restarts after long (several-week) intervals of missing data.

#### Technical Details

Details for the separate regression and EWMA methods are given in the preceding pages. The adjusted R^2^ coefficient for the regression is tested each day. This coefficient does not give the quality of regression but is employed here specifically as a measure of daily predictive capability using an empirically derived threshold criterion. When the data pass this test, the model is assumed to have explanatory value, and the regression algorithm is applied. When the data fail this test, the EWMA algorithm is used. The Poisson distribution test is applied when less than a week (3-6 days) of recent data is available. A Poisson distribution is assumed with mean and variance equal to the mean of the recent counts. An alert is issued if the current count exceeds this mean and if probability that the current count was drawn from this distribution is less than 1% (red alert) or 5% (yellow alert). Practical safeguards for the composite method are as described in the regression and EWMA sections above.(19)

#### Benefits

This algorithm is the default because it is designed to avoid mismatching the method to the data. The regression model accounts for the expected data trends when they are seen in the baseline. When they are absent because of the case definition used to filter the data, because of the size of the monitored region, or because of data problems, alerting is based on the EWMA algorithm.

#### Limitations

The goodness-of-fit test occasionally misclassifies the data. The test is set to err toward the more conservative EWMA to avoid misfitting the data model.

### Algorithm: C1, C2, and C3

#### Categorization

Adaptive Control Chart

#### Purposes

To purpose is to detect general data aberrations. Algorithms C1, C2, and C3 of the Early Aberration Reporting System (EARS) developed at the Centers for Disease Control and Prevention are used in many U.S. states and in numerous foreign countries.(24) They are included in the ESSENCE suite because of their wide application. While they lack many of the features described above, their simplicity has both benefits and limitations.

#### Technical Details

The C1 algorithm subtracts the daily count from the mean of a moving baseline ending the previous day. In effect, it then divides this difference by the standard deviation of counts in that baseline. If the result exceeds 3, indicating an increase above the mean of more than 3 standard deviations, an alert is issued. The C2 algorithm does the same calculation but imposes a 2-day buffer between the test day and the baseline. The C3 algorithm is a more sensitive version of C2 that adds the values from the 2 previous days if they do not exceed the threshold. All three algorithms use the same criterion of an increase of at least 3 baseline standard deviations above the sliding baseline mean. An important implementation detail is that ESSENCE does not use the standard 7-day baseline because substantial experience has shown that for many time series, such a short baseline gives an unstable statistic that can lead to a loss of confidence in the results. The implemented baseline is 28 days as in the EWMA and regression methods.(21) There are no other changes to the standard EARS methods, including retention of the flat 3-standard-deviation threshold regardless of the data stream.

#### Benefits

The methods are easy to understand and widely known.(25–27)

#### Limitations

Like the EWMA, the methods take no account of systematic data behavior such as day-of-week effects or seasonal trends. C3 is the only one of these methods with sensitivity to gradual outbreak effects, but it is known to produce high alarm rates. For all three methods, threshold data values for alerting may fluctuate noticeably from day to day.

### Summary Alerts--adjustment for multiple testing

#### Categorization

False Discovery Rate processing of multiple alerts

#### Purpose

The *parallel monitoring problem* is the monitoring of multiple separate time series representing different physical locations, such as counties or treatment facilities, possibly stratified by other covariates such as syndrome type or age group. The purpose of the Summary Alert Algorithm is to maintain sensitivity while limiting the number of alerts that arise from testing the numerous resulting time series.

Multiple testing can lead to uncontrolled alert rates as the number of data streams increases. For example, suppose that a hypothesis test is conducted on a time series of daily diagnoses of influenza-like illness. In a one-sided test, this test results in a statistic whose value in some distribution yields a probability p that the current count is as large as observed. For a desired Type I error probability of ***α***, the probability is then (1−*α*α that an alert will not occur in the distribution assumed for background data. Thus, for the parallel monitoring problem of interest here, if such tests are applied to N independent data streams, the probability that no background alerts occur is (1− *α*^*N*^, which decreases quickly for practical error rates ***α*** For a single-test error rate of ***α*** = 0.05, for example, the probability of at least one background alert exceeds 0.5 if more than 13 independent tests are applied.

#### Technical Details

For N tests, where N is the number of combinations of region, syndrome, age group, and any other covariates affecting the number of tests, let *P*_(1)_,…, *P*_(*N*)_ be the *p*-values sorted in ascending order, an ordering that puts the smallest and most significant p-value first. The Summary Alert method applies the Simes-Seeger-Eklund criterion to reject the combined null hypothesis of no anomaly for any series.(28) The null hypothesis is rejected if for some *j*\*, *j*\* = 1,..,N, *P*_(*j*\*)_ < *j*\**α* / *N*. To interpret this condition, note that for the most significant p-value, an alert requires that *P*_(*1*)_ < *α/N* the strict Bonferroni bound. If *α* = 0.01 and N = 50, then the condition becomes *P*_(*1*)_ < 0.0002. For the least significant p-value, the condition is simply *P*_(*N*)_ < *α* highly unlikely for the weakest result.

If this condition is satisfied for any j*, then test results are considered alerts for all j < j*.(29) The Summary Alert is implemented at two levels, FDR and FDR-Major. For the FDR level applied to N time series, the implementation is as above. For a more liberal option appropriate for certain syndromes or scenarios, FDR-Major applies the condition to two sets of N/2 time series.

#### Benefits

In defining the false discovery rate as the expected ratio of false alerts to the total alert count, Benjamini and Hochberg showed that the Simes-Seeger-Eklund criterion gives an overall error rate of *α* if the N time series tested are statistically independent.(30) Overall, this criterion avoids the excess alerting resulting from using the nominal threshold *α* for all data streams and also avoids the loss of sensitivity from using only the Bonferroni bound *α/N*.

#### Limitations

If one of the p-values crosses the adjusted threshold, it is not obvious for epidemiological or other reasons which tests to consider anomalous. Most users have followed the natural procedure described by Simes to consider all p-values less than *P*_(*j*\*)_ as individual alerts. Another limitation is that in general the time series are not statistically independent. For situations where dependence is known, Hommel recommended the condition *P*_(*j*)_ < *j* ∙ i / *C* ∙ *N* where *C* = Σ 1/*j*. In ESSENCE applications where many groups of time series may be requested and dependence can change, the above condition with C = 1 is applied.

### Spatial cluster determination

#### Categorization

Spatial Scan Statistics

#### Purpose

A problem with sophisticated temporal detectors is choosing the appropriate size and location of the collection region for time series counts. If this region is too small or mislocated, cases may be missed and the baseline data may not have enough structure, but if the region is too large, the scale and variability of the large-scale time series may reduce sensitivity by masking clusters of interest. We apply spatiotemporal scan statistics in an attempt to promptly localize public health problems. For ESSENCE, JHU/APL built and implemented a Java version of the algorithm implemented in the SaTScan software of Martin Kulldorff originally developed for spatial surveillance of cancer and subsequently used and enhanced for many types of hotspot detection.(31)

#### Technical Details

The null hypothesis is that the set of data subregions (often zip codes) in the recent time interval tested forms a random sample from an expected spatial distribution of cases. The expected distribution is not uniform over subregions but reflects a “customary” spatial case spread that reflects urban/suburban case ratios or other factors. ESSENCE implementation calculates the expected spatial distribution using recent case counts from a sliding baseline interval. In effect, the code is similar to a common application of SaTScan, the space-time permutation scan statistic, restricted to test cases from only the most recent time interval and assuming circular clusters.(32)

As in SaTScan, the method calculates a test statistic for each candidate cluster. The test statistic in the ESSENCE implementation is Kulldorff’s Poisson log likelihood ratio. The set of candidate clusters is generated by scanning over a set of cluster center locations, often taken as centroids of all zip codes in the dataset, and considering all circles within a maximum radius of each center, where the number of circles is limited by the number of data subregions within each radius. The maximum test statistic over these candidates is then tested for significance. Statistical significance inference does not depend on a theoretical distribution but on repeated trials on simulated datasets randomly drawn using the baseline distribution. For each such trial, the algorithm uses the same scanning procedure to derive a trial maximum.

For assessing the significance of the maximum test statistic over all observed clusters, the ESSENCE code uses the Gumbel distribution method.(33) The code collects 99 trial maxima, fits a Gumbel distribution to these values, and uses the fitted distribution to assign a p-value to the test statistics of clusters found in the original data. The observed cluster with the maximum test statistic is considered significant if its p-value is below a predetermined threshold, often set to 0.01. This threshold criterion can yield multiple significant clusters in a given run if more than one candidate cluster yields a test statistic whose p-value is below the threshold. For each significant case cluster, the system shows the location, extent, and degree of significance using the GIS software.

#### Benefits

The ESSENCE Java implementation inherits features that have popularized SaTScan. Potential clusters of interest are localized without bias regarding the center or extent of the cluster as well as the spatial resolution of the data allows. As noted in Kulldorff, Heffernan, et al., the empirical significance testing with many repeated trials takes “into account the multiple testing stemming from the many potential cluster locations and sizes evaluated.”(32)

#### Limitations

The most important limitation, applicable also to SaTScan and to all other spatial or space-time cluster detection methods, is that the usefulness of the method strongly depends on the reliability of the expected spatial distribution. The use of census-based distributions, insurance eligibility lists, regression models, and other means have been used to derive the expected distribution. The method implemented in ESSENCE infers this distribution from recent data separated from the test date(s) by a 2-day buffer.

Evaluation of statistically significant clusters for epidemiological significance is a nontrivial task which may be exacerbated if the number of significant clusters is misleading or excessive because the expected distribution is unrepresentative or because investigation resources are insufficient.

The use of this popular approach for prospective use has been criticized despite numerous applications and published real-life successes,(34) and the ESSENCE implementation lacks the prospective adjustment in SaTScan attempting to manage cluster rates for multiple successive runs. The ESSENCE implementation also does not support elliptical cluster shapes, simultaneous clustering of multiple data sources, or test statistics other than the Poisson log likelihood ratio. The user with a sufficiently detailed dataset and an application that requires these extended SaTScan features should be aware of these limitations.

### Time-of-arrival aberration detection

#### Categorization

Multiple Automated Hypothesis tests

#### Purpose

This algorithmic approach was implemented to find and display unusual clusters of syndromically related emergency department visits by patients arriving for care within a short time interval.

#### Technical Details

Patient visit counts are tabulated by cells, with one cell for each hospital/time-interval/sub-syndrome combination. See Figure 1.

**Figure 1:**
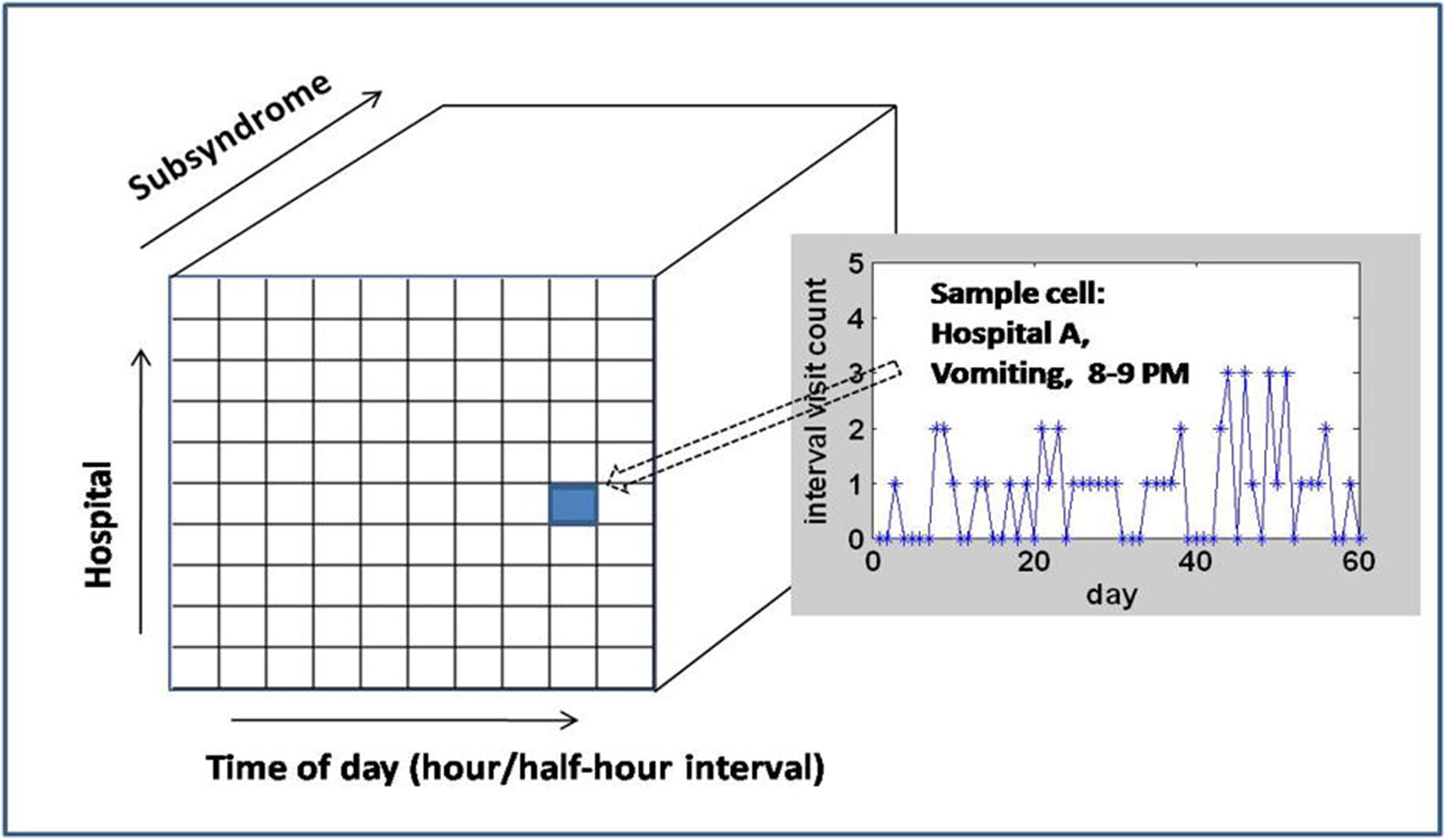
Schematic of time-of-arrival filtering and anomaly detection process

- For the visit counts in each cell, a Poisson or negative binomial test is chosen using the last 60 days of visit counts for that cell. The Poisson distribution is used unless the count variance exceeds the mean by a factor of 1.1 or greater, and then the time series is considered overdispersed. This situation occurs for relatively few cells, generally corresponding to the more common (sub)syndromes for the largest hospitals at the busiest times when most alerts would be generated. For this situation, a negative binomial distribution is assumed. Parameter settings and distribution types were derived from testing on several years of patient visit records from a variety of large and small hospitals.
- Once the distribution is chosen, parameters for each cell are calculated from the 60-day baseline. For each cell, an alert is then flagged if the current count exceeds the upper limit threshold for the chosen distribution based on a preselected p-value.
- Based on empirical results using 12 years of data from 134 hospital EDs from a large state with labeled events, a threshold p-value of p* = 10-4 (0.0001) was chosen.
- Time intervals for the cells are 30 min., 60 min. beginning on the hour, and 60 min. beginning on the half hour, again a result of empirical testing.
- Practical overrides are implemented based on observed cell counts. At least three observed cases are required for an alert. This minimum may be increased for more common syndromes. Mandatory alerts may also be implemented for certain subsyndrome/count combinations, such as subsyndromes for severe illness, regardless of the hypothesis test.

#### Benefits

In validation testing to monitor visit clusters for 51 subsyndromes for 134 hospitals at the time intervals above with the chosen p-value threshold, alert rates were consistently manageable and found all known clusters from a small historical collection of events except for two groups of 3-4 visits at very busy times. The alert burden was still manageable at the county level when anomalous clusters for all hospitals within each county were combined. The simplicity of this approach allows multiple daily runs and adaptation to new improvised subsyndromes with rapid system response without impact on routine processing.

#### Limitations

The hypothesis tests include no direct modeling of seasonality or other systematic data behavior. They were implemented to enable county-level processing, and validation was conducted on a 12-year historical dataset from one state. Expanding the computational load to include much larger sets of hospitals or syndrome groups with limited investigation capability may require recalibration (p-value threshold, minimum alert counts) or an alternate approach to retain sensitivity with manageable alerting.

### Identification of term-based free-text anomalies

#### Categorization

Hypothesis test for excessive occurrence of selected chief complaint terms

#### Purpose

This algorithm was implemented to point out unusual distributions of chief complaint terms of interest without dependence on syndrome definitions.

#### Technical Details

For each term in any chief complaint in the most recent time interval, the number of chief complaints containing that term in this interval is tested relative to the number of chief complaints containing same term in a much larger sliding baseline interval, assumed to be representative of customary data.

- The algorithm forms 2×2 contingency tables whose entries are:

A = number of recent chief complaints with the term,
B = number of recent chief complaints without the term,
C = number of baseline chief complaints with the term,
D = number of baseline chief complaints without the term
- For many such tables with small cell counts, Fisher’s Exact Test is applied to alert when the probability that count of the term of interest is ≥ A, given the contingency table’s marginal totals, is below a critical p-value.
- If the smaller of B and C is larger than 1000, then a chi-square test is applied to the same contingency table with negligible loss of accuracy. The critical threshold is then applied to half the resultant (two-sided) p-value to determine anomalous terms.
- Results are shown only for candidate terms that have not been previously eliminated because they are stopwords with no informational content (such as “the”, “if”, “all”) or because users have previously added them to a list of terms to be ignored (such as “patient”, “complaint”, “test”).
- Conventions adopted from empirical test results and also to avoid impacting ESSENCE processing are: The test interval is 24 hours, the baseline interval is 30 days, a buffer of 7 days is implemented between test interval and baseline, and the threshold p-value is set at p* = 10^−5^ (0.00001).

#### Benefits

With the above p-value threshold and settings, this method detects as few as three instances of unusual terms (place names, rare signs/symptoms) and unusual concentrations of interesting common terms while averaging from 0-4 anomalous terms per day over small and large hospitals. Inspecting chief complaints containing each of a small number of terms each day and disqualifying some terms from further consideration is a manageable task that can uncover clusters of visits that could be missed by syndromic methods. In testing with historic data, chief complaints containing anomalous terms found with the strict p-value threshold adopted have included small clusters of visits resulting from documented events of food poisoning and heat-related illness. This analysis has also found new abbreviations used by hospitals in their chief complaints. These new abbreviations can then be added to the Chief Complaint Processor to improve syndrome and subsyndrome categorization.

#### Limitations

While the simplicity of this method avoids impact on daily ESSENCE processing or user investigation of large collections of distributed data streams, the method has no means to interpret anomalous terms, identify phrases with multiple terms, or distinguish topics of concern in chief complaints that may not share specific terms. The only preprocessing of free-text terms is the application of ESSENCE Chief Complaint Processor spell checking and abbreviation/acronym expansion. Interpretation of the anomalies requires a human-in-the-loop, both to evaluate anomalous terms for investigation and to rule-out specific terms from future anomalies. The method is subject to effects of changing terminology (street drug names, triage vocabulary and abbreviations), and the user should be aware of current perceived health threats and corresponding emergency/urgent care language.

